# Feasibility and Acceptability of the Multicontext Approach for Individuals with Acquired Brain Injury in Acute Inpatient Rehabilitation

**DOI:** 10.1101/2020.05.10.20064261

**Authors:** Abhishek Jaywant, Chelsea Steinberg, Alyson Lee, Joan Toglia

## Abstract

The Multicontext (MC) approach, a metacognitive intervention designed to improve awareness, strategy use, and executive functioning, may be beneficial for individuals with acquired brain injury (ABI) undergoing acute inpatient rehabilitation. The goal of this study was to provide evidence of feasibility, acceptability, and patient perceived benefit of the MC approach. A case series of eight individuals with acquired brain injury and at least mild executive functioning impairment were recruited from an acute inpatient rehabilitation unit. The MC approach— involving guided questioning and patient self-generation of strategies within everyday functional cognitive tasks—was implemented within routine occupational therapy. Outcome measures were self-report of treatment satisfaction, the Self-Regulation Skills Interview, and the Weekly Calendar Planning Activity. Participants rated the MC approach as highly satisfying and engaging. They described subjective improvements in their ability to use executive functioning strategies. The MC approach was associated with improvement in awareness, strategy use, and executive functioning at the conclusion of treatment. The MC approach may be a beneficial structured intervention for individuals with acquired brain injury and executive dysfunction undergoing acute inpatient rehabilitation. Further evaluation with larger samples in controlled trials is warranted.

## Introduction

Executive functions—including the ability to organize information, maintain it in working memory, inhibit distractions, and shift to relevant aspects of the task— are important cognitive skills for the efficient performance of adaptive, goal-directed everyday activities [1,2]. Following acquired brain injury (ABI)—such as stroke, traumatic brain injury, and brain tumor—executive functioning skills are frequently compromised [3–5]. Compounding these impairments, persons with ABI frequently experience poor awareness of their cognitive deficits [6,7] and lack insight into the importance of using cognitive strategies to facilitate their performance in everyday activities.

Metacognitive strategy training is a cognitive rehabilitation technique that has demonstrated efficacy in the post-acute phase of brain injury. Metacognitive techniques focus on improving ABI individuals’ awareness, self-monitoring, and ability to initiate and implement effective cognitive strategies to facilitate goal-directed behavior. The Cognitive Rehabilitation Task Force (CTRF) of the American Congress of Rehabilitation Medicine recently published its updated guidelines on cognitive rehabilitation for ABI. It recommended metacognitive strategy training as a practice standard for the post-acute phase following ABI, specifically for improving attention and mild-to-moderate executive dysfunction [8]. However, the CTRF guidelines also noted that cognitive rehabilitation interventions have varying evidence for their ability to generalize learned skills to everyday activities [9,10].

Acute inpatient rehabilitation is a critical phase of recovery. Cognitive treatments delivered at this phase of recovery can capitalize on a critical period of brain plasticity [11]. Unfortunately, there is relatively limited evidence supporting the use of cognitive rehabilitation techniques such as metacognitive strategy training during this phase of rehabilitation. Further, usual care in rehabilitation tends to be focused on physical impairments and recovery and thus persons with ABI may not have the opportunity to recognize difficulties in higher-level cognitively-based instrumental activities of daily living. This may result in persistent cognitive impairment [12] and negatively impact return to work and other occupations. Conversely, intervening during the acute rehabilitation period can increase the potential of individuals with ABI to return to their everyday activities sooner and at a greater independence level after discharge from the hospital. In small studies of acute inpatients with stroke, strategy training outside of routine care has demonstrated preliminary evidence for feasibility [13], improved awareness and strategy use [14], and improved cognitive flexibility and inhibition on neuropsychological assessment [15]. Metacognitive training has also demonstrated feasibility in inpatient psychiatric care [16].

The Multicontext (MC) approach is one form of metacognitive strategy training that may be well-suited to the acute inpatient rehabilitation setting. The MC approach [17–19] was developed to help persons with ABI increase their awareness of cognitive performance and ability to effectively apply executive functioning strategies across a range of functional tasks and everyday activities. This is important because many individuals with ABI have difficulty connecting similarities across activity experiences, including failing to recognize that similar cognitive symptoms are hindering performance across situations. The MC approach structures treatment activities in a “horizontal” manner by presenting a series of activities that gradually differ in surface features or physical characteristics, while requiring similar cognitive demands. For example, a patient may be presented with tasks that require maintaining three items in working memory, which is practiced across activities such as placing appointments in a daily schedule, organizing a pillbox, and searching for items on a menu. A therapist guides the patient to become aware of performance errors and self-generate cognitive strategies.

The MC approach has previously demonstrated feasibility and efficacy in outpatient treatment in individuals with traumatic brain injury [20], Parkinson’s disease [21], and multiple sclerosis [22]. To date, however, it is unknown whether the MC approach can be integrated within routine clinical care in the acute inpatient rehabilitation setting for individuals with ABI. Given that the goals of inpatient rehabilitation are to return patients as close as possible to their prior level of independence in everyday activities, the MC approach may be a form of metacognitive treatment that is especially well-suited to this setting, facilitating cognitive performance in functional activities such as medication management, scheduling appointments, bill paying, and cooking that patients may otherwise not have the opportunity to practice in a therapeutic setting in the acute post-injury phase.

The goal of this study was to evaluate the feasibility, acceptability, and patient satisfaction of the MC approach in a series of eight cases with ABI and executive dysfunction within the acute inpatient rehabilitation setting. Patient satisfaction and perceived benefit are important metrics in developing and refining interventions because of their importance for patient motivation and engagement [23]. We hypothesized that the MC approach could be integrated within routine rehabilitation care and that patients who complete the MC approach would find the treatment to be engaging, beneficial, and perceived as helpful in improving their functional cognitive skills. A second, exploratory goal was to provide data on improvements in awareness, strategy use, and executive functioning at the conclusion of treatment.

## Materials and Methods

### Participants

Participants were eight consecutively-recruited individuals with ABI recruited from an acute inpatient rehabilitation unit (IRU) at a large, urban academic medical center. All procedures were ethically approved by the medical center’s Institutional Review Board. Inclusion criteria included: age between 18 and 80 years; English-speaking; confirmed diagnosis of ABI based on radiological assessment; able to comprehend multistep directions and participate in conversation as assessed by the Functional Independence Measure (FIM; required score of 4 or above on Comprehension and Expression items); impaired performance (<2 SD below demographically-corrected normative data) on at least one screening measure of executive functioning; able to attend to a cognitive task for at least 10 minutes; cognitively independent in basic self-care activities; able to read standard size newsprint; and the ability to demonstrate functional use of at least one hand. Patients who were admitted to the IRU and who met inclusion/exclusion criteria were identified by the occupational therapy supervisor on admission, who described the project and obtained written consent.

Table 1 provides baseline demographic scores for each individual in the case series, labeled P1 through P8. Four individuals experienced a stroke, two were diagnosed with a brain tumor, one experienced a traumatic brain injury, and one was diagnosed with a primary central nervous system cancer. All participants were admitted to the IRU and provided informed consent to participate in the study. The majority of patients were in the acute phase following injury, though P1 had a relatively longer time since onset of injury.

### Screening Assessments

The Montreal Cognitive Assessment (MoCA) [24] is a 30-point performance-based cognitive screening instrument that is a standard of care measure on our IRU. It assesses the domains of visuospatial/executive function, attention, language, abstraction, memory, and orientation. A higher score indicates better cognitive performance. The standard cutoff for cognitive impairment is a score less than 26/30. The Trail Making Test (TMT) and Symbol-Digit Modalities Test (SDMT) were used to screen for executive dysfunction. The TMT is a neuropsychological measure of visual attention and processing speed (TMT-A) as well as rapid attentional shifting and cognitive flexibility (TMT-B). The SDMT [25] is a timed assessment of divided attention, working memory, incidental learning, and psychomotor speed. As depicted in Table 1, all participants demonstrated impaired performance on at least one of the TMT-A, TMT-B, and SDMT.

### Outcome Assessments

#### Patient Satisfaction Questionnaire

At the conclusion of treatment, participants were given a questionnaire that was developed for this study by the authors (see Supplemental Material). Participants were asked open-ended questions of what they liked most and least about the program, what they would have changed about the program, and any additional suggestions or recommendations. This questionnaire also included Likert-type ratings of their satisfaction, enjoyment, perceived benefit from the intervention, and perceived likelihood of continuing to use the strategies learned in treatment.

#### Self-Regulation Skills Interview (SRSI)

The SRSI [26] is a clinician-administered semi-structured interview that assesses an individual’s metacognitive skills and ability to use cognitive strategies. It comprises six questions and each question is scored on a 10-point Likert-type scale with lower scores indicating greater metacognitive skills (i.e., lower scores are desirable). It has demonstrated strong interrater reliability and test-retest reliability in individuals with ABI [26]. It consists of a total score and three factors, Awareness, Readiness to Change, and Strategy Behavior. We administered the SRSI prior to and after the MC intervention and were particularly interested in the Total Score (0-60) as well as the Awareness (0-20) and Strategy Behavior (0-30) subscores, as these have previously been shown to be diminished in individuals with ABI [26].

Examples of interview questions include “Can you tell me how you know that you experience [cognitive difficulty]; that is, what do you notice about yourself?” (Awareness subscale) and “What strategies are you currently using to cope with your [cognitive difficulty]?” (Strategy Behavior subscale).

#### Weekly Calendar Planning Activity (WCPA)

The WCPA [27,28] is a standardized, performance-based, ecologically-valid measure of executive functioning in which the participant has to organize a list of appointments into a weekly schedule. Effective performance requires the individual to plan an effective approach, maintain in mind multiple task rules, problem-solve, avoid conflicts, and inhibit distracting information. The outcome variable was the percentage of appointments entered correctly relative to the total number of appointments entered.

#### Functional Independence Measure (FIM)

The FIM [29] is a standard-of-care measure of disability that assesses the level of assistance required for an individual to perform activities of daily living. The FIM is comprised of 18 items that assess bathing, grooming, eating, upper/lower body dressing, toileting, bowel/bladder management, transfers, locomotion, stair mobility, comprehension, expression, social interaction, problem-solving, and memory. Each item is rated on a 1-7 scale, with higher scores indicating greater functional independence. The FIM Total was used in this study, with scores ranging from 18 to 126.

### Intervention: the Multicontext (MC) Approach

The MC approach has been described in detail previously (see [17–20] for a full description). In brief, the MC approach used in this study involved 30-minute sessions of strategy and self-monitoring practice within the context of everyday activities. It was determined that a minimum of 6 structured activities across a minimum of 3 sessions would be used. Following the 3^rd^ session, activities that were chosen by the client could be used.

The initial focus of MC treatment is on helping the individual to self-discover error patterns and learn to anticipate cognitive performance challenges through repeated structured experiences, across functionally relevant activities. Activities can address the person’s motor goals while simultaneously requiring targeted cognitive performance skills (e.g., searching and locating information, keeping track, or shifting between task steps and directions). For example, activities and materials such as a schedule or menu can be presented on a bulletin board or wall with lists or cards placed on a table or another part of the room to require standing, reaching and/or walking while completing the activity. Self-check lists are incorporated across treatment activities so that the person can self-evaluate their own work and self-discover performance errors themselves, with guided questions as necessary. The focus is not on awareness of deficits per se but on awareness of performance [30]. Awareness of “performance” includes appraisal of task difficulty, recognition of performance challenges, identification of task methods that contributed to success (versus those that did not) and accurate evaluation of performance outcomes. A metacognitive framework of guided questions before, during and after each activity is utilized.

Pre-made activity kits that included directions and everyday materials such as menus, schedules, coupons or food circulars were available, along with guidelines for using and positioning the materials. The activities involved use of functional contexts such as locating items from a list on a food circular or menu, gathering items on a list from a supply closet or highlighting items on a list, or scheduling appointments on a calendar according to specific criteria. Therapists were able to choose from directions that were consistent across different functional activities and that placed similar demands on targeted areas such as working memory, flexibility, inhibition, planning and organization, depending on the client’s cognitive symptoms, performance errors and needs. In addition, the activities could be easily adjusted to the person’s cognitive and motor abilities, completed within 10-15 minutes and included pre-made answer checklists so that the patient could self-check their own work with guidance [31].

One of two occupational therapists delivered the intervention. Guided questions were used throughout treatment to facilitate error detection or monitoring and self-assessment of performance. Guided learning methods are similar to use of Socratic questioning as described by others (see for example [32] for use in cognitive therapy for depression). Prior to the task, the therapist asked questions to help participants anticipate challenges and to self-generate executive functioning strategies [17,19]. Therapists would ask, for example, “What kind of challenges do you anticipate having during this task?” and “What special strategies or methods could you use to complete everything you have to do?” The participant would then attempt the task while the therapist observed participant performance. If necessary, the therapist would mediate in the middle of the task to probe the participant’s self-assessment of success or challenge. After the task was complete, the therapist used guided questioning to ask participants what errors they made, what strategies they used, what strategy modifications or alternatives could be used in the future (“What could you do differently next time?”), and what other activities this same strategy would apply to including activities that had been completed in-session previously (“What other activities would using a checklist be helpful for?”). The latter question was used to facilitate transfer across practiced and non-practiced activities. Therapists also utilized praise and encouragement to reinforce participant success.

### Procedure

Occupational therapists implementing the MC approach were trained and supervised by the senior author. This included attendance at inservices, readings, review of case videos, and weekly meetings to review cases. A minimum of 3 treatment sessions were videotaped and reviewed by the senior author using a fidelity checklist to insure adherence to the protocol [19].

Participants were administered the cognitive screening measures and the pre-treatment SRSI and WCPA by their treating occupational therapist (OT). The treating OT was not blind to these assessments because they required this assessment data to tailor the MC intervention to the specific participant. Each participant completed between six and nine consecutive MC activities within their routine occupational therapy session in inpatient rehabilitation. Physical goals were simultaneously addressed by choosing or arranging activities in ways that required motor skills such as dexterity, reaching, standing or walking. Patients were seen twice a day during their rehabilitation stay and MC treatment was delivered either daily or twice a day. Session lengths were approximately 30 minutes. The total number of activities/sessions varied depending on the participant’s length of stay. Post-treatment follow-up SRSI, WCPA, and satisfaction questionnaire were completed by a separate therapist who was blind to the participant’s initial assessment scores and their progress in treatment.

### Data Analysis

We used both descriptive/qualitative and inferential/quantitative approaches in our statistical analyses. To evaluate participant satisfaction, we qualitatively describe the responses to the post-treatment patient satisfaction questionnaire. We provide two narratives from participants P5 and P7 to qualitatively illustrate the MC approach and describe within-session observations. These narratives were extracted by a neuropsychologist (AJ) who reviewed session videotapes and who was not involved in the assessment or treatment of individual participants. Graphical analysis was conducted for each participant to evaluate the magnitude of change on each of the pre/post assessment measures. Given the small sample size and single group pre/post design, we conducted exploratory within-subjects nonparametric testing (Wilcoxon signed-rank tests) to determine the statistical significance of changes in assessment measures at the group level. Median and interquartile range (IQR) are provided for these group-level analyses. Effect sizes were calculated using Cohen’s d for paired samples.

**Table 1.** Demographic and clinical information for each participant.

|  | **Age Range** | **Gender** | **Pre-Injury Functioning** | **Diagnosis** | **Time Since Onset** | **Length of Treatment (Days)** |
| --- | --- | --- | --- | --- | --- | --- |
| **P1** | 70-79 | F | Retired, independent | Bifrontal Brain Tumor | 4 weeks | 8 |
| **P2** | 40-49 | F | Working, independent | TBI and subarachnoid hemorrhage | 10 days | 8 |
| **P3** | 60-69 | F | Retired, independent | Left frontoparietal Stroke | 7 days | 8 |
| **P4** | 30-39 | F | Working, independent | Cancer with multiple hemorrhagic lesions | 6 days | 6 |
| **P5** | 60-69 | F | Working, independent | L hemisphere Stroke | 8 days | 7 |
| **P6** | 60-69 | F | Retired, independent | L hemisphere Stroke | 4 days | 6 |
| **P7** | 20-29 | F | Working, independent | Left frontal Brain Tumor | 5 days | 8 |
| **P8** | 70-79 | M | Working, independent | Right hemisphere Stroke | 12 days | 8 |

*Note.* We provide age ranges and only general details regarding pre-injury occupational functioning to protect participant confidentiality.

**Table 2.** Clinical screening results for each participant.

|  | **MoCA Total Score (/30)** | **TMT-A age-corrected z-score** | **TMT-B age-corrected z-score** | **SDMT age-corrected z-score** |
| --- | --- | --- | --- | --- |
| **P1** | 18 | -- | -- | -- |
| **P2** | 24 | -6.85 | -4.21 | -2.36 |
| **P3** | 15 | -3.29 | -5.68 (2 errors) | -2.09 |
| **P4** | 25 | -3.35 | -3.16 (1 error) | -3.00 |
| **P5** | 21 | -9.09 | -5.68 (4 errors) | -3.92 |
| **P6** | 23 | -6.45 | -5.68 (2 errors) | -0.20 |
| **P7** | 25 | +0.49 | -10.14 (5 errors) | -2.09 |
| **P8** | 28 | -1.11 | -0.01 (1 error) | -2.10 |

*Note.* MoCA = Montreal Cognitive Assessment; SDMT = Symbol Digit Modalities Test; TBI = traumatic brain injury; TMT = Trail Making Test. Participant 1 was not administered the TMT or SDMT at admission to the rehabilitation unit.

## Results

### Demographic Characteristics

Demographics characteristics and medical information is provided in Table 1. Four patients had stroke, two had primary brain tumors, one had metastatic cancer, and one had a traumatic brain injury. All were functionally independent prior to onset of illness and several were working. Participants’ median age was 64.5 years (IQR = 36-71.5 years). Our sample was well-educated: one individual had a high school education, two had college degrees, and five had graduate degrees. The median time since onset of brain injury was 7.5 days (IQR = 5.25-11.5 days). Median length of treatment including OT initial evaluation and discharge evaluation was 8 days (IQR = 6.25, 8.0). As depicted in Table 2, median score on the MoCA was 23.5, below clinical cutoff for impairment (IQR = 18.75-25). Participants demonstrated significant impairment in baseline screening measures of executive functioning including the TMT-A, TMT-B, and SDMT.

### Client Satisfaction

On the client satisfaction questionnaire, all but one participant reported that their strategy use improved “very much” or “extremely.” All participants described finding the MC intervention “very much” or “extremely” satisfying, and the majority of participants endorsed “a lot” of enjoyment. Participants stated that the aspects that they liked least were the time commitment and additional work required, emotions elicited (i.e., frustration), and challenges at the outset of the treatment. Despite these challenges, all participants derived subjective benefit from the program, noting increased awareness (“helped me think”), practicality of the strategies learned (“simplicity of strategies”), and benefit from working on everyday activities that were relevant to their lives and goals (“Real life activities, not abstract tasks”; “Things were practical, gave me a sense of where I was at.”). No participant experienced adverse events.

### Patient Narratives of Treatment

We describe narratives and ratings from two participants. P5 was a stroke survivor in her 60s. Her initial treatment goals were to return to work and driving, and to improve her word-finding. During an initial MC treatment session in which she had to schedule appointments into a calendar, she described minimal pre-task challenges of having “limited time”, and articulated in a very limited sense one possible strategy she could use of “classifying by category.” After the task, she again had difficulty articulating use of a strategy as she stated, “I probably was using strategies but I don’t remember them.” She described the task as only “slightly” challenging. On a subsequent task that required her to create a schedule of leisure activities, she again identified the task as only minimally challenging, though she did more clearly articulate a strategy prior to the task (“breaking the task down”). Post-task, she noted that it was also important for her to check her work for errors. During a later task requiring her to organize items in a mock kitchen, the participant clearly articulated two strategies to assist her in working memory and short-term maintenance of information (“categorizing” and “repeating things [to myself], especially things that are unusual because they may be harder to remember”). She successfully executed this strategy by grouping together items that belonged within a certain category (e.g., spices). She was also much more aware of her errors, describing the task as challenging and acknowledging that she “missed some items.”

**Table 3.** Patient initial goals, structured activities, and strategies.

|  | **Initial Treatment Goals** | **Example Multicontext Treatment Activities** | **Strategies Emphasized** | **Awareness of Performance and/or Strategy Use at Discharge** |
| --- | --- | --- | --- | --- |
| **P1** | “To be able to drive a car, go back to living in [state], and participate in social activities.” | Keeping track of items on a list and shifting attention, using schedules, composing an email, ordering food online, reading and summarizing an article, looking at grocery coupons | Creating a plan prior to a task, self-checking during and after a task. | Acknowledges some cognitive difficulties, though others are minimized. “I am hesitant to drive.” “My son will have to help me with walking the dog.” |
| **P2** | Return home and live independently<br>Return to work | Searching for, organizing, and selecting information in a shopping catalog, airline ticket, online drugstore, Excel spreadsheet, and menu | Planning ahead, breaking tasks down, Underlining/markings relevant information | “I am slower in paper tasks and things requiring more organization.”<br>“Use of colors helps me sort and organize. Repeating is helpful [for memory].” |
| **P3** | “I just want to be the way I used to be.” | Shifting between 2-3 stimuli and following written directions. Motor activity (clothespins), laundry, card games, menus, medication pillbox | Use finger to block out distractions, verbal rehearsal, breaking down and chunking information, double checking work | “I have difficulty with memory when there is a lot of information at once.”<br>Identifies using her finger and breaking items down to one component at a time and states “they work very well.” |
| **P4** | “I want to go back to work. I want to go back to cooking and doing things at home without needing assistance.” | Searching and keeping track of information in pictures of grocery store items, retrieving kitchen ingredients, bills, pillbox organizer, cooking | Verbal rehearsal, writing lists and checking items off, double-checking accuracy | “My mind wanders and I can tell I’m off focus. [I have difficulty with] paying bills, keeping track of medicine, and other activities with several steps.”<br>Identifies strategies of rehearsal and writing information down, notes she has to use them more often. |
| **P5** | “Return to work and driving.” | Searching, locating, and keeping track of information in schedules, on the computer, in the kitchen, and in a card game | Categorizing, chunking, verbal rehearsal, taking pauses | “I need to pay attention. I can’t remember certain things like words.”<br>“I need extra time to think.” Identifies categorizing, chunking, and rehearsal as helpful strategies. |
| **P6** | “To walk. To be able to write better.” | Searching for and highlighting items on a list and placing them in different locations. Brunch menu, recreation/activity schedules, food pantry, motor activities | Using finger/ card for stimuli reduction (blocking out); verbal rehearsal, breaking down and chunking information, double checking work | “It is still frustrating because it is not easy as it was before but now I take time to think and concentrate. I know I need to repeat things to myself and chunk things together.” |
| **P7** | “Get better and return to work.” | Search and locate using a coupon book, a schedule of films, online shopping, and investigating exercise classes online | Pre-planning, Verbal rehearsal, self-talk | “I am more organized in my approach. I know I have to plan and structure my day” |
| **P8** | Balance, walking, fine motor skills:<br>“To get dressed by myself” | Searching, keeping track of and shifting between stimuli using calendar, brunch menu, news articles online, online shopping | Taking time before jumping into activities and monitoring speed. Circling key details/words or most important points. Double-checking. | Takes time to review tasks and accurately identifies challenges. Identifies priorities within complex tasks before starting. Able to monitor when he needs to slow down and when he needs to take a break due to mental fatigue. Consistently double checks work. Self-identified the need to schedule activities that require more concentration earlier in the day. |

P7 was in her 20s and had been diagnosed with a brain tumor. Her treatment goals were to return to work and get better physically in terms of her balance and fine motor skills. Her first activity was to find exercise classes at certain times that would fit a mock schedule. She did not acknowledge any potential challenges or the need to use any strategies. She began the task quickly. Some awareness emerged at the end of the task, as she allowed that she was “a little slow” and (with prompting from the therapist) stated that she “could have taken notes.” On a subsequent task in which she had to organize the schedule for a film festival, she demonstrated increasing awareness of the need to use strategies prior to the task. However, her description of the strategy was vague. After attempting the task, with prompting from the therapist, the participant was able to see that she had been using strategies to facilitate her performance—these included starring certain films and crossing off others. To facilitate transfer, the therapist helped the participant see that her approach could be useful in managing different kinds of schedules as well as webinars. At the end of treatment, P7 completed an online shopping task. She clearly articulated strategies pre-task, including “taking notes” and “marking it up… highlighting things I need to remember.” She completed the task without errors and while doing so, generated an additional strategy that could be helpful in the future “breaking up the task.” P7 became increasingly adept at combining multiple strategies to facilitate her performance.

### Exploratory Analysis of Clinical Outcome

As shown in Figures 1 and 2, all participants demonstrated an improvement on the SRSI Total Score, Awareness subscale, Strategy Use subscale, and WCPA from pre-treatment to post-treatment. Group statistics are provided in Table 2. As a group, there was a statistically significant improvement from pre-treatment to post-treatment in SRSI Total Score, SRSI Awareness subscore, SRSI Strategy Use subscore, WCPA, and the FIM. Table 3 provides participants’ goals, treatment activities used, and narrative responses to the SRSI at discharge. All participants were able to articulate cognitive strategies, many of which entailed strategies to better plan/organize, keep track of information, and self-check work for errors.

**Figure 1.**
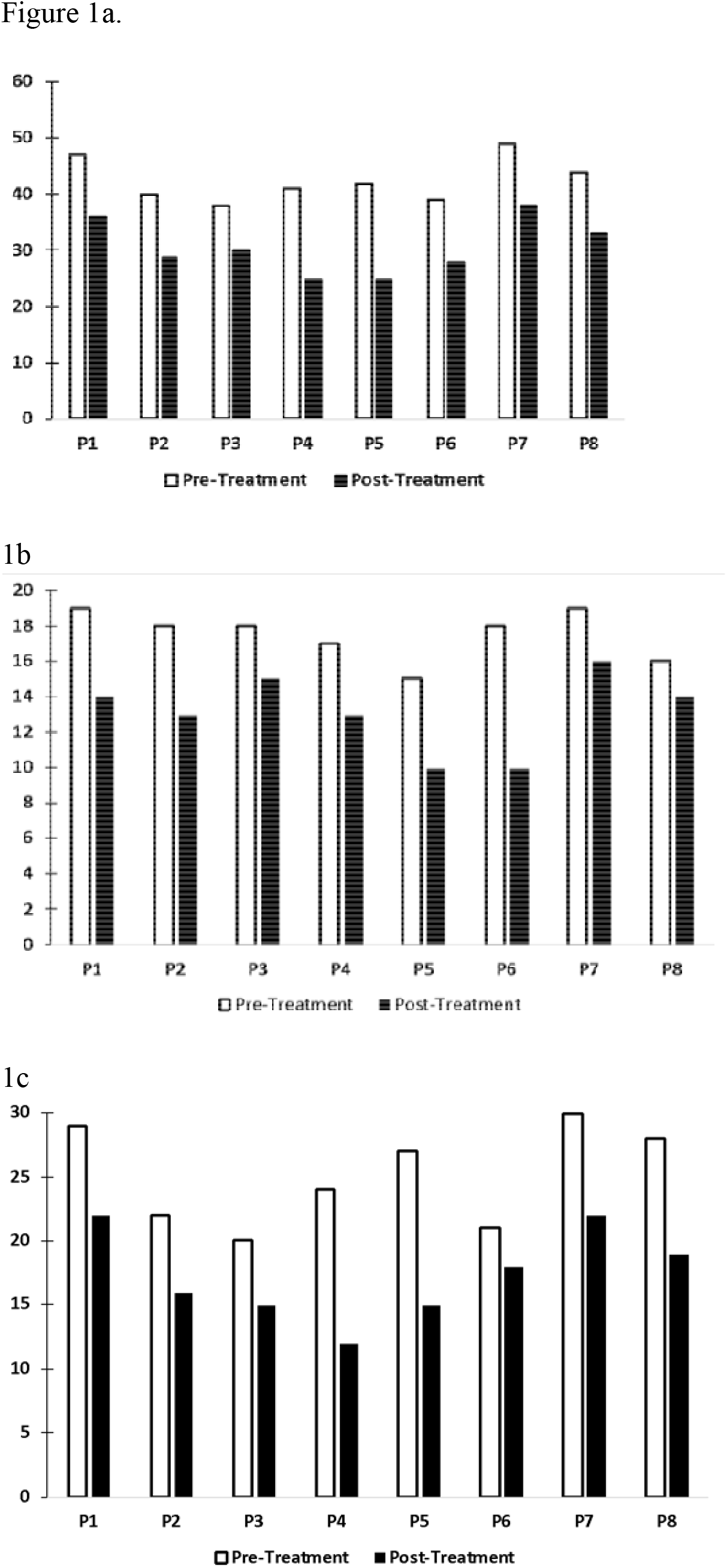
Pre-treatment to post-treatment change on the Self-Regulation Skills Interview (SRSI) (a) Total Score, (b) Awareness subscale, and (c) Strategy Use subscale. Lower scores on the SRSI denote better awareness, strategy use, and self-regulation.

**Figure 2.**
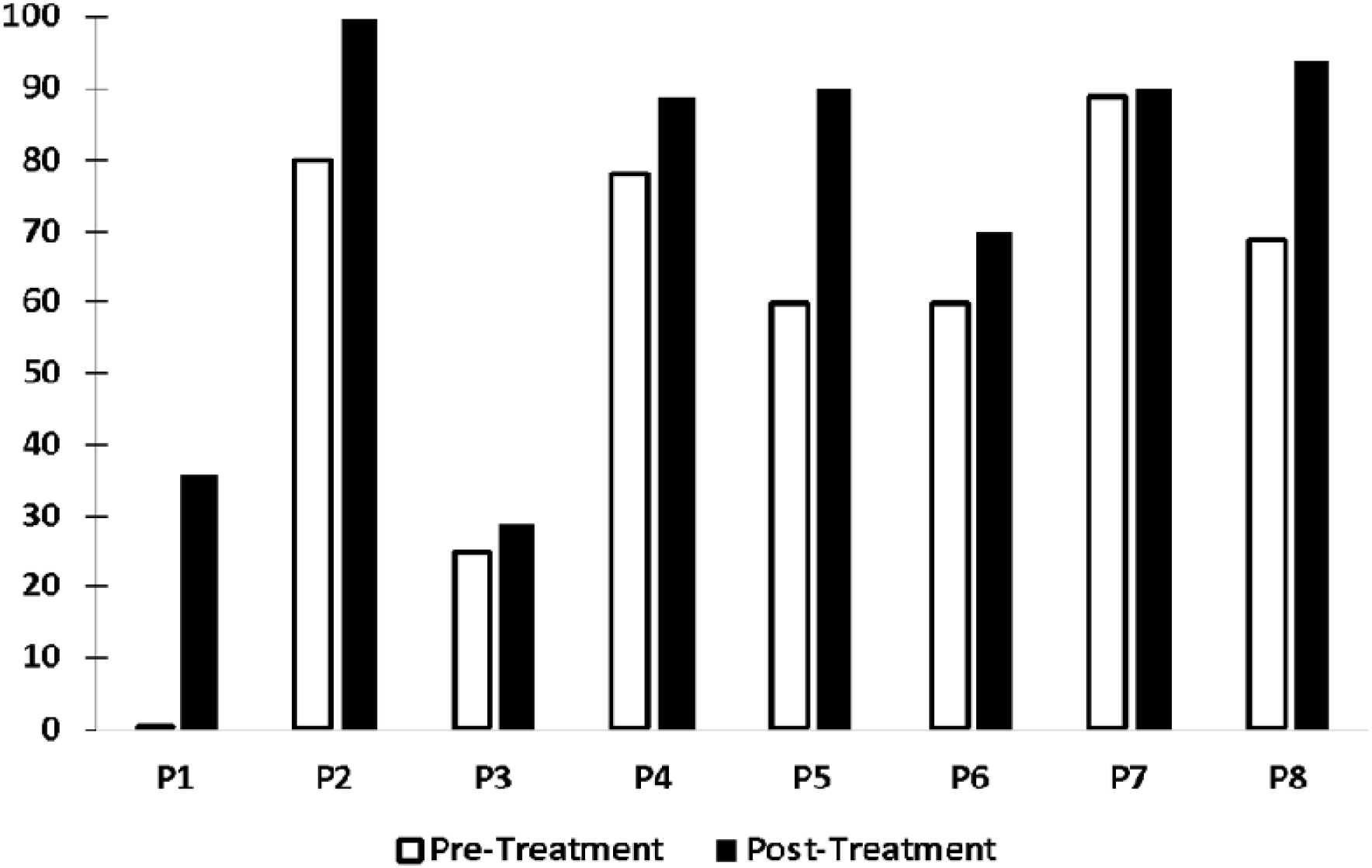
Pre-treatment to post-treatment change on the Weekly Calendar Planning Activity (WCPA). The outcome measure is the percentage of accurate appointments entered into the calendar, calculated by dividing the number of appointments accurately entered by the total number of appointments entered. A higher percentage correct indicates better performance.

**Table 4.** Summary of outcome measures.

|  | **Pre-Treatment**<br>**Median** | **Pre-Treatment**<br>**IQR** | **Post-Treatment**<br>**Median** | **Post-Treatment**<br>**IQR** | **Wilcoxon Z** | **p-value** | **Cohen's d** |
| --- | --- | --- | --- | --- | --- | --- | --- |
| **SRSI Total** | 41.5 | 39.25-46.25 | 29.5 | 25.75-35.25 | 2.59 | .01 | 4.07 |
| **SRSI Awareness** | 18 | 16.25-18.75 | 13.5 | 10.75-14.75 | 2.54 | .01 | 2.37 |
| **SRSI Strategy Use** | 25.5 | 21.25-28.75 | 17 | 15-21.25 | 2.52 | .01 | 2.42 |
| **WCPA** | 64.5 | 33.75-79.5 | 89.5 | 44.5-93 | 2.52 | .01 | 1.36 |
| **FIM** | 80.5 | 74.25, 87 | 96 | 91.5-99 | 2.52 | .01 | 1.62 |

*Note.* FIM = Functional Independence Measure; IQR = Interquartile Range; SRSI = Self-Regulation Skills Interview; WCPA = Weekly Calendar Planning Activity.

## Discussion

The primary goal of this study was to evaluate the feasibility, acceptability, and patient satisfaction of the MC approach in a series of eight cases with ABI and executive dysfunction within the acute inpatient rehabilitation setting. A second, exploratory goal was to investigate improvements in awareness, strategy use, and executive functioning at the conclusion of treatment. Our principal finding is that participants rated the intervention as highly satisfying and described subjective improvements in their awareness of and ability to use executive functioning strategies. Despite the relatively short treatment durations and the time constraints of the acute inpatient rehabilitation setting, we found that the MC approach integrated within usual care may improve awareness, strategy use, and executive functioning.

Ascertaining client engagement and satisfaction was the primary goal of this study. At the conclusion of treatment, we evaluated participants’ subjective impressions of the intervention. All participants reported that they found the MC treatment to be enjoyable, satisfying, and beneficial. Qualitatively, we observed that as participants experienced challenges in structured activities, they began to identify additional cognitively-based IADL activities that they wanted to try and were able to articulate new, cognitively-based goals. All but one participant rated their ability to apply compensatory strategies as significantly improved. These descriptions indicate that participants were engaged and motivated to participate in the intervention and suggests that further study of the MC approach is warranted. This was the first study to evaluate MC treatment within routine care in the acute inpatient setting. It is notable that patients found the treatment highly satisfying despite the frequent comorbid medical management required in this setting and despite the significant physical and cognitive demands of standard rehabilitation therapies.

A unique aspect of the MC approach is that it directly and explicitly attempts to facilitate transfer to untrained but related everyday activities. Therapists attempted to facilitate transfer and generalization by specifically guiding participants to see links between various tasks. This transfer was highlighted in P7’s narrative, whereby she took a strategy of highlighting key elements of task instructions in one task (scheduling) and applied it to another task (online shopping). She was able to articulate that these strategies could be helpful for managing various schedules and webinars. As demonstrated in Table 3, at discharge all participants articulated clear and well-defined challenges and barriers to their cognitive performance as well as strategies that could help them manage the cognitive demands of everyday tasks. These strategies comprised rehearsal, categorization, writing information down, annotating stimuli to keep track of key details, and self-checking work for errors.

The eight individuals with ABI treated with the MC approach demonstrated significant improvements in awareness, strategy use, and executive functioning. This was apparent despite the fact that qualitatively, our sample all initially identified fully or in-part only physically-based goals. This may have been enhanced by the integration of cognitive and motor activities during the early part of treatment. Functional cognitive activities were presented in a manner that simultaneously addressed the person’s physical goals, such as standing balance, walking or reaching, particularly during initial sessions.

As described in our patient narratives and as measured by the SRSI, patients’ awareness of their errors and of the importance of strategy use emerged during treatment Through task practice and therapists’ guided questioning, participants generated strategies to facilitate working memory, planning, and organization and attempted to use them in personally relevant, simulated tasks. From pre-treatment to post-treatment, participants’ organization and planning improved on an objective, standardized measure of working memory and executive functioning (WCPA). Our results add to a large body of research that has demonstrated positive effects of experiential practice, guided questioning, and metacognitive strategy training for brain injury in the post-acute phase of recovery [8,33]. Importantly, we extend this work to the acute inpatient rehabilitation setting, and complement recent studies demonstrating positive effect in acute settings [16,34,35]. Our results indicate that the MC approach can be integrated within routine therapy—with pre-made activity kits [31], guidelines for therapist fidelity, and therapist training and feedback— with positive benefits for awareness and strategy use in persons with ABI.

This study had several limitations, including the small sample size and lack of control group. We sought to provide preliminary feasibility and acceptability data to inform future controlled clinical trials with larger sample sizes. The inclusion of a control group is important given that spontaneous neurologic recovery often occurs during the acute period following brain injury and likely impacted the size of our treatment effects. Another limitation is that variability in participant lengths of stay resulted in a different number of sessions for participants. However, this limitation reflects the realities of implementing interventions within acute inpatient medical units. Further, while all participants had a form of brain injury affecting executive functioning, the heterogeneity of diagnoses within the category of acquired brain injury may mask important differences in treatment efficacy and satisfaction for different neurologic conditions. Given the time constraints of the inpatient setting, we administered only two neuropsychological measures of executive functioning. Further study of the MC approach would benefit from a more comprehensive neuropsychological battery to evaluate the effects of the treatment on specific aspects of executive functions. We also did not have long-term follow-up to examine intervention effects over time, or additional assessments of transfer to everyday activities, which is important to investigate in future research.

## Conclusion

In this case series of eight individuals with ABI, we demonstrated that in the acute inpatient rehabilitation setting, individuals with ABI found the treatment to be satisfying and engaging, and perceived subjective improvement in their ability to use cognitive strategies to facilitate performance in everyday activities. Further, the MC approach was associated with significant improvements in awareness, strategy use, and executive functioning. Despite frequently not having cognitive goals at the outset of treatment, participants were engaged in functional cognitive activities, began to recognize cognitive challenges, and acknowledged the importance of using cognitive strategies. Goal revision and adjustment during early stages of intervention may be an indicator of increased awareness and should be further explored in future investigations. These findings suggest that larger clinical trials of the MC approach are warranted. Based on additional studies with larger sample sizes and appropriate control groups, the MC approach may be a useful adjuvant treatment for individuals with brain injury and executive dysfunction during acute rehabilitation.

## Data Availability

Data are not publicly available but may be obtained via a request to the authors.

## Acknowledgements

The authors thank Andrea Mastrogiovanni MA, OTR/L and Dan Tufaro MA, OTR/L for their assistance with this project, and the clients for their participation and contributions to this study. We also thank Michel O’Dell MD, for his support with this study.

